# Steroid Hormones in Dementia: A Cross-Diagnostic Molecular Analysis of Blood and Cerebrospinal Fluid

**DOI:** 10.64898/2026.02.12.26346149

**Authors:** Tik Muk, Asger Wretlind, Kourosh Hooshmand, Marc Clos-Garcia, Liu Yue, Anja Hviid Simonsen, Laura M. Winchester, Tarun veer S. Ahluwalia, Petroula Proitsi, Riccardo E. Marioni, Thomas Kümler, Steen Gregers Hasselbalch, Cristina Legido-Quigley

## Abstract

**Introduction:** Alzheimer’s disease (AD) disproportionately affects women, with accumulating evidence suggestion a contributary role of hormones in this disparity. Given the known influence of hormones on brain health and cognition, characterizing specific profiles in dementia is crucial. In addition, sex-stratified hormonal alterations in AD and other dementias remain poorly understood.

**Methods:** We quantified nine steroid hormones: 11-deoxycortisol, 17-hydroxyprogesterone, aldosterone, cortisol, dihydrotestosterone, estrone, progesterone, testosterone and estradiol. The hormones were quantified in cerebrospinal fluid (CSF) and plasma from 204 participants across five cognitive categories: no cognitive impairment (n=32), mild cognitive impairment (MCI) non-AD (n=38), MCI due to AD (n=21), AD dementia (n=81), and vascular dementia (VaD) (n=32). Participants were recruited at the Danish Dementia Research Centre, Copenhagen University Hospital, Copenhagen, Denmark. Hormone levels were measured using liquid chromatography-tandem mass spectrometry. Sex-stratified generalized linear models were adjusted for age. Overall, 50.5% of participants were women with a mean age of 69 (SD = 9.8) compared to men with a mean age of 70 (SD = 9.1).

**Results:** In women with AD, CSF cortisol and 11-deoxycortisol were significantly elevated compared to women with no cognitive impairment (Fold Change (FC) (95% CI) = 1.13 (1.01-1.27), p-value = 0.04 and (FC (95% CI) = 1.01, (1.00-1.01), p-value = 0.03, respectively). Plasma progesterone was decreased (FC (95% CI) = 0.90 (0.81, 0.99), p-value = 0.04). Women with VaD exhibited reduced CSF estradiol (FC (95% CI) = 0.86 (0.74, 0.98), p-value = 0.03). In men with AD, plasma aldosterone was elevated (FC (95% CI) = 1.19 (1.06, 1.33), p-value = 2.81e-03).

Correlation analyses revealed that CSF cortisol in women was significantly correlated with CSF AD pathology markers in amyloid-beta 42 (r = -0.29, p-value = 3.02e-03) and phosphorylated tau (r = 0.2, p-value = 0.04). The increase of cortisol was validated in an external cohort where t-test showed significant difference in cortisol between people with AD and cognitively healthy controls (CN), this difference was larger in women (mean AD = 0.26 vs mean CN = 0.21, p-value = 1.79e-06) than men (mean AD = 0.23 vs mean CN 0.21, p-value = 0.04)

**Conclusion:** Our findings demonstrate sex-dependent dysregulation of steroid hormone in dementia. Specifically, cortisol and aldosterone are highlighted, which are potential modifiable targets.

## Introduction

Alzheimer’s disease (AD) is a debilitating neurological condition and the primary cause of dementia, with a significant gender gap projected to widen further over the coming decades ^1^. Epidemiological studies have shown that women are approximately one and a half times more likely to develop AD compared to men, particularly above the age of 80 ^2,3^. One potential factor contributing to the difference in AD risk between men and women is their differing exposure to hormones ^4^.

The relationship between endogenous sex hormone exposure, in the form of the natural, cumulative presence of estrogens (including estrone, estradiol, and estriol), progesterone and testosterone, and dementia risk in women have shown inconclusive findings. Some studies have found that reproductive events related to shorter endogenous sex hormone exposure, such as early menopause, are associated with higher risk of all-cause dementia ^5,6^ and poorer cognitive outcomes ^7,8^. Further studies from the UK Biobank demonstrated that women experiencing early or surgical menopause, thereby curtailing their natural hormone exposure, showed a heightened risk of all-cause dementia, whereas prolonged exposure appeared protective, particularly among those without the apolipoprotein E ε4 (*APOE* ε4) allele ^9,10^. However, contradictory evidence exists, with studies reporting that longer endogenous hormone exposure was associated with accelerated brain aging ^11^, amyloid-beta 42 and p-tau levels ^12^.

In men, lower levels of testosterone have been consistently associated with all-cause dementia and AD ^13–15^. It is also worth noting that steroid hormones like cortisol and aldosterone have been implicated in AD pathology. Cortisol levels are elevated in both mild cognitive impairment (MCI) and dementia due to AD compared to healthy controls ^16,17^, and high cortisol levels are associated with cognitive decline and higher AD risk ^18–20^. For aldosterone, evidence is more limited. However, one study found higher aldosterone concentration associated with impaired cognitive function ^21^, while another study investigating individuals with primary aldosteronism revealed an increased risk for dementia in this population ^22^.

Collectively, these clinical studies suggest a role for steroid hormones in cognitive decline and dementia risk. In this study, we measured in paired cerebrospinal fluid (CSF) and plasma steroid hormone profiles in individuals with dementia subtypes to further understand the relationship between hormonal regulation and dementia pathology.

## Methods

### Participants

This study included 204 participants recruited from the Danish Dementia Research Centre at Copenhagen University Hospital, Rigshospitalet, following written informed consent and in accordance with the Declaration of Helsinki. Patients were diagnosed after comprehensive evaluation^23^ and categorized into five distinct dementia subgroups defined by their clinical syndrome and etiological diagnosis. All patients had measures for CSF peptide biomarkers amyloid-β 42 (Aβ-42), phosphorylated tau (pTau) and total tau (tTau), as part of the diagnostic routine. The groups were: participants with no cognitive impairment (NCI) (subjective impairment) (n = 32), participants with MCI-AD (n = 21), participants with MCI and no AD pathology: MCI non-AD (n = 38), participants with AD dementia (n = 81), and participants with vascular dementia (VaD) (n = 32). Plasma and CSF samples were stored at -80°C until analysed.

### Targeted hormone panel

Steroid hormones were extracted from plasma and CSF using the same method, a full description of this steroid quantification method can be found in the supplementary material section S1. In short, 50 μL of each sample were extracted using acetone:acetonitrile (1:1 v/v) protein precipitation, followed by centrifugation, solvent evaporation, and reconstitution in methanol:water (1:1 v/v). Hormone analysis was performed using liquid chromatography-tandem mass spectrometry (LC-MS) on an Agilent 1290 UHPLC system coupled to an Agilent 6460 triple quadrupole mass spectrometer. Chromatographic separation was achieved using a Waters HSS T3 column (1.8 μm, 2.1 × 100 mm) using a binary gradient system: water containing 2 mM ammonium fluoride (mobile phase A) and methanol as mobile phase B. Detection was performed using electrospray ionization in positive mode with dynamic multiple reaction monitoring (dMRM). Each hormone was quantified using external calibration curves with specific standards (0.00075-1000 ppb, 21 points). Data processing and peak integration were performed using Agilent MassHunter Quantitative Analysis Software (version 10.2).

### Statistical Analyses

Demographic and clinical characteristics were re-analyzed in the subset of participants included in the steroid hormone assessment as part of this secondary analysis. Data distribution was evaluated prior to the statistical analysis using the Shapiro–Wilk test for normality and Breusch–Pagan for homoscedasticity. Non-normally distributed variables were log_2_-transformed prior to analysis. Group comparisons were conducted by comparing every cognitive impairment category to a prespecified reference group (no cognitive impairment, NCI), using linear models with treatment-versus-control contrasts. Analyses were performed in R using the packages emmeans (v1.11.0) for marginal means and contrasts, sandwich (v 3.1.0) for HC3 covariance estimation, lmtest (v0.9.40) for Breusch–Pagan testing, and tidyverse for data structuring and management.

Steroid hormones measures with more than 30% missing values across samples were excluded. One participant from the no cognitive impairment group was identified as an outlier and excluded due to abnormally high plasma concentrations of progesterone and 17-hydroxyprogesterone, resulting in a total of 204 participants. Principal component analysis (PCA), variable importance in projection (VIP) score analysis, and partial least squares– discriminant analysis (PLS-DA) were applied to evaluate potential group separations across cognitive impairment categories.

Group differences in individual steroid hormones were examined both within each sex and in the combined sexes using generalized linear models (GLMs). Models were adjusted for relevant clinical covariates, including age at visit, sex and the interaction between group and sex (sex was included only for combined-sex analyses). Fold changes (2^β) with 95% confidence intervals (CIs) and corresponding p-values were visualized using forest plots generated with ggplot2 (v3.5.1). The concentrations of steroid hormones were summarized as mean with standard error of the mean (SEM). A p-value < 0.05 was considered statistically significant. Correction for multiple testing was carried out with FDR adjustment.

Associations between steroid hormones and Alzheimer’s disease–related clinical characteristics were assessed using Spearman’s rank correlation, with and without sex stratification. Correlation coefficients and corresponding p-values were visualized in a Z-score–normalized heatmap generated using the corrplot package in R. All statistical analyses and data visualizations were conducted using R Studio software (version 4.3.0; Boston, MA, USA).

### Cortisol validation

Data from the Alzheimer’s Disease Neuroimaging Initiative (ADNI)^24^ was used as an external validation of measures of cortisol, a total of n = 426 participants where included, 286 categorized as cognitively healthy and 140 individuals had AD. AD was assessed by neurologic examinations and standardized neuropsychological assessments. Cortisol was measured in plasma with a targeted LC-MS platform, using a Shimadzu LC system coupled to a SCIEX QTRAP mass spectrometer. Separation was achieved on an Acquity UPLC BEH C18 column (Waters). Comparison between levels of cortisol in cognitively healthy and individuals with AD was carried out with t-tests on log-transformed normalized data.

## Results

### Participant characteristics

This study examined hormone levels in plasma and CSF from 204 participants with varying degrees of cognitive decline. The overall mean age was 69 (SD = 9.5) years. Participants with no cognitive impairment were youngest, mean age 65 (SD = 10.4) years, while those with VaD were oldest with a mean age 75 (SD = 6.5) years. The MCI, MCI Non-AD, and AD groups had mean ages of 66 (SD = 7.4), 68 (SD = 11.5), and 70 (SD = 8.3) years, respectively. Approximately half the participants were women (50.5%, n = 103), though sex distribution varied across groups. The group with the highest percent of women was the AD group which included 62.9% women (n=51), while the group with the lowest percent of women were the MCI Non-AD group had 26.3% women (n=10) (Table 1). The overall mean age of women was 69 (SD = 9.8) compared to men with a mean age of 70 (SD = 9.1).

**Table 1.** Participant Characteristics.

|  | Overall | P-Value (To NCI)* |  |  |  |
| --- | --- | --- | --- | --- | --- |
| n | 204 | MCI | MCI Non-AD | VaD | AD |
| Age (y) | 69.45 ± 0.67 | 0.95 | 0.56 | <0.001 | 0.07 |
| Aβ42 (pg/ml) | 616.90 ± 22.10 | <0.001 | 0.78 | <0.001 | <0.001 |
| pTau (pg/ml) | 59.69 ± 2.03 | 0.002 | 0.81 | <0.001 | <0.001 |
| tTau (pg/ml) | 453.04 ± 19.71 | <0.001 | 0.19 | <0.001 | <0.001 |
| MMSE Score | 24.97 ± 0.33 | 0.57 | 0.02 | <0.001 | <0.001 |
| Female |  |  |  |  |  |
|  | NCI | MCI | MCI Non-AD | VaD | AD |
| n | 18 | 9 | 10 | 15 | 51 |
| Age (y) | 65.83 ± 2.78 | 67.11 ± 2.86 | 62.10 ± 3.44 | 75.60 ± 2.07 | 69.53 ± 1.21 |
| Aβ42 (pg/ml) | 870.61 ± 66.87 | 448.67 ± 26.07 | 1075.50 ± 104.49 | 655.93 ± 98.64 | 450.74 ± 10.98 |
| pTau (pg/ml) | 40.65 ± 3.02 | 48.67 ± 6.90 | 41.50 ± 3.11 | 85.85 ± 7.92 | 76.68 ± 4.12 |
| tTau (pg/ml) | 244.83 ± 21.23 | 342.11 ± 50.34 | 274.70 ± 28.07 | 696.29 ± 79.32 | 621.82 ± 38.60 |
| MMSE Score | 28.06 ± 0.79 | 27.56 ± 0.47 | 26.14 ± 0.88 | 23.15 ± 1.17 | 23.07 ± 0.74 |
| Male |  |  |  |  |  |
|  | NCI | MCI | MCI Non-AD | VaD | AD |
| n | 14 | 12 | 28 | 17 | 30 |
| Age (y) | 64.29 ± 2.50 | 65.58 ± 1.95 | 70.61 ± 2.09 | 75.35 ± 1.20 | 71.10 ± 1.43 |
| Aβ42 (pg/ml) | 1005.57 ± 65.43 | 505.67 ± 72.65 | 829.26 ± 57.66 | 391.93 ± 35.20 | 405.50 ± 15.54 |
| pTau (pg/ml) | 39.00 ± 3.25 | 78.58 ± 8.02 | 43.81 ± 3.25 | 47.20 ± 5.26 | 64.32 ± 5.10 |
| tTau (pg/ml) | 230.43 ± 21.67 | 524.67 ± 64.83 | 312.23 ± 41.90 | 356.64 ± 51.03 | 518.13 ± 54.39 |
| MMSE Score | 28.54 ± 0.61 | 27.17 ± 0.67 | 26.00 ± 0.65 | 22.00 ± 1.41 | 23.68 ± 0.78 |

Data displayed as mean ± SEM
\*All p-values are computed using combined-sex data and compared each class to the No Cognitive Impairment (NCI) reference group.

### Hormone characteristics

We quantified nine unique steroid hormones across plasma and CSF samples. Eight hormones were quantified in plasma, while five were quantified in CSF. Four hormones: aldosterone, 11-deoxycortisol, cortisol, and testosterone, were successfully measured in both sample types, whereas estradiol was only detectable in CSF. Hormone concentrations spanned a 100-fold range. Testosterone in CSF showed the lowest mean concentration at 0.12 ng/ml in NCI men and 0.09 ng/ml for NCI, while cortisol in plasma had the highest mean concentration at 69.15 ng/ml NCI men and 74.52 NCI women. Several hormones exhibited differing concentrations between men and women, notably dihydrotestosterone being detectable solely in the plasma of men.

### Hormones associated with AD

Sex-stratified generalized linear models adjusted for age revealed distinct hormonal patterns in CSF and plasma across diagnostic groups. In CSF (Figure 1, Supplementary table 1), women with AD exhibited significantly elevated levels of 11-deoxycortisol and cortisol (Fold change (FC) (95% CI) = 1.01, (1.00-1.01), p-value = 0.03 and FC (95% CI) = 1.13 (1.01-1.27), p-value = 0.04, respectively), while women with VaD showed decreased estradiol levels (FC (95% CI) = 0.86, (0.74-0.98), p-value = 0.03). No significant associations between CSF hormones and cognitive impairment were observed in men.

**Figure 1.**
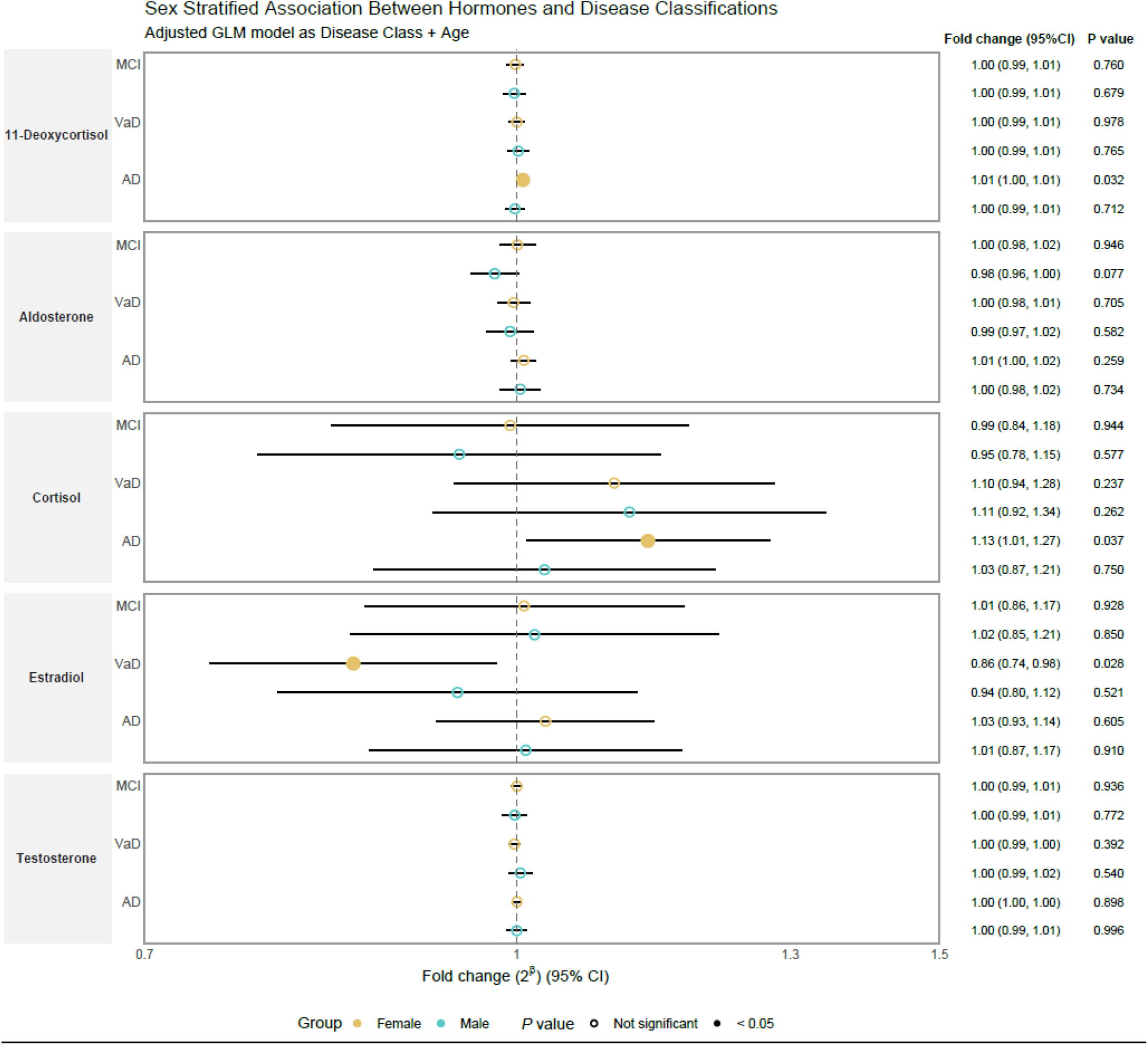
CSF Sex stratified GLM. Forest plot of association between CSF hormones and disease classification using GLM. Fold change with 95% confidence interval against NCI and p-values are shown on the right.

Plasma hormone analyses revealed sex-specific patterns (Figure 2). In women, lower progesterone showed a significant association with AD (FC (95% CI) = 0.90, (0.81-0.99), p-value = 0.04). In men, elevated 17-hydroxyprogesterone was associated with MCI (FC (95% CI) = 1.08, (1.00-1.15), p-value = 0.04), but not with AD or VaD. Elevated estrone was associated with VaD in men (FC (95% CI) = 1.45, (1.05-1.99), p-value = 0.03). Additionally, higher plasma aldosterone was associated with AD in men (FC (95% CI) = 1.19 (1.06-1.33) p-value = 2.81e-03), this was the only finding that remained significant after FDR correction (q = 0.02).

**Figure 2.**
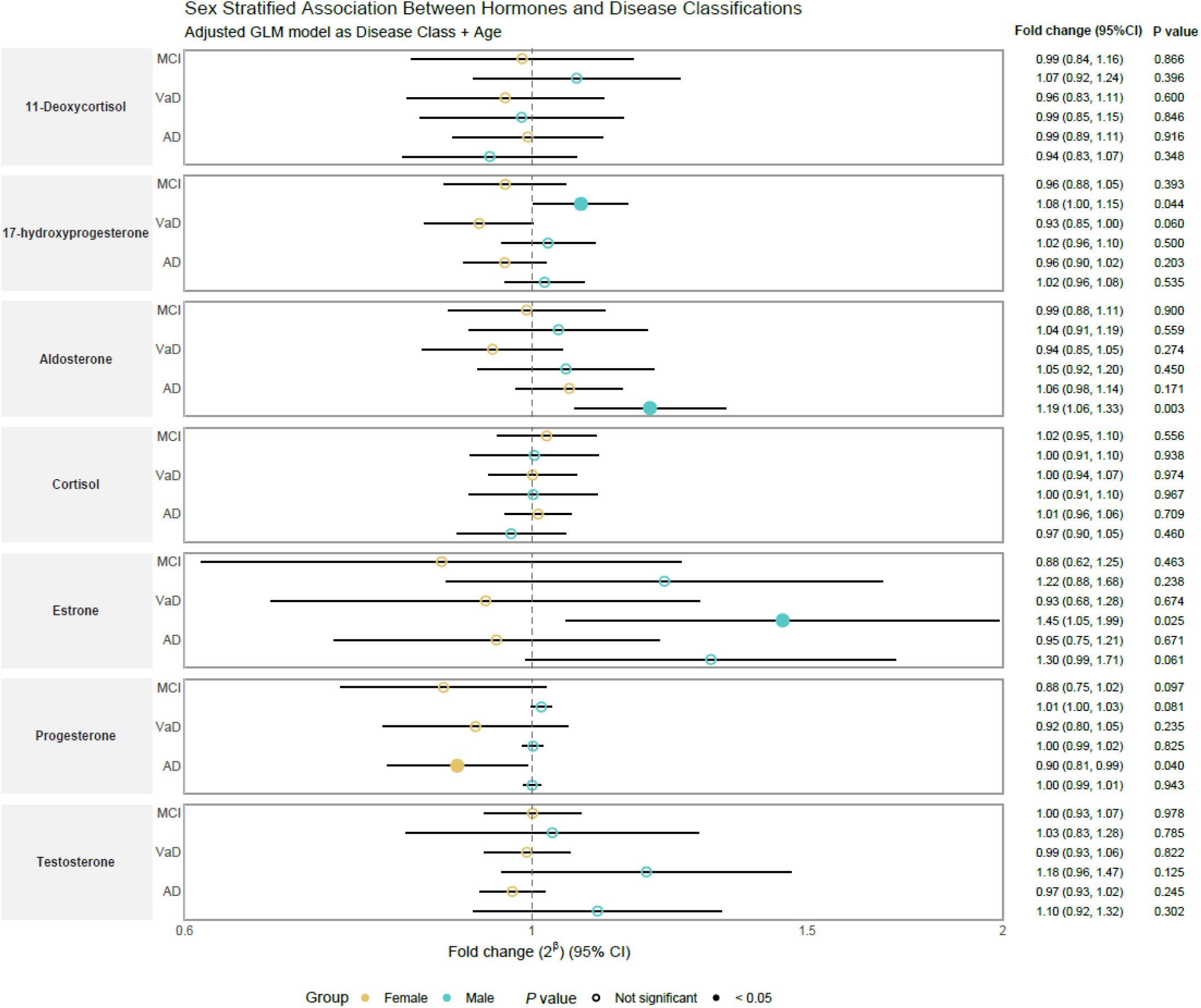
Plasma Sex stratified GLM. Forest plot of association between plasma hormones and disease classification using GLM. Fold change with 95% confidence interval against NCI and p-values are shown on the right.

Sex-adjusted generalized linear models were also conducted. CSF cortisol was significantly elevated in individuals with AD compared to NCI (FC (95% CI) = 1.50, (1.03-2.18), p-value = 0.03; Supplementary Figure 1). Plasma progesterone was significantly lower across all cognitive impairment groups compared to controls: MCI (FC (95% CI) = 0.88, (0.78-0.98), p-value = 0.02), AD (FC (95% CI) = 0.89, (0.82-0.96), p-value = 2.01e-03, q = 0.02), and VaD (FC (95% CI) = 0.9, (0.82-0.99), p-value = 0.03; Supplementary Figure 2). Furthermore, plasma 17-hydroxyprogesterone was significantly decreased in individuals with VaD compared to controls (FC (95% CI) = 0.91, (0.83-1), p-value = 0.04).

### Hormone correlations

Spearman correlation analyses revealed correlations between hormones, AD biomarkers, and cognitive function, with notable sex-specific patterns (Figure 3, Supplementary Figures 3 and 4).

**Figure 3.**
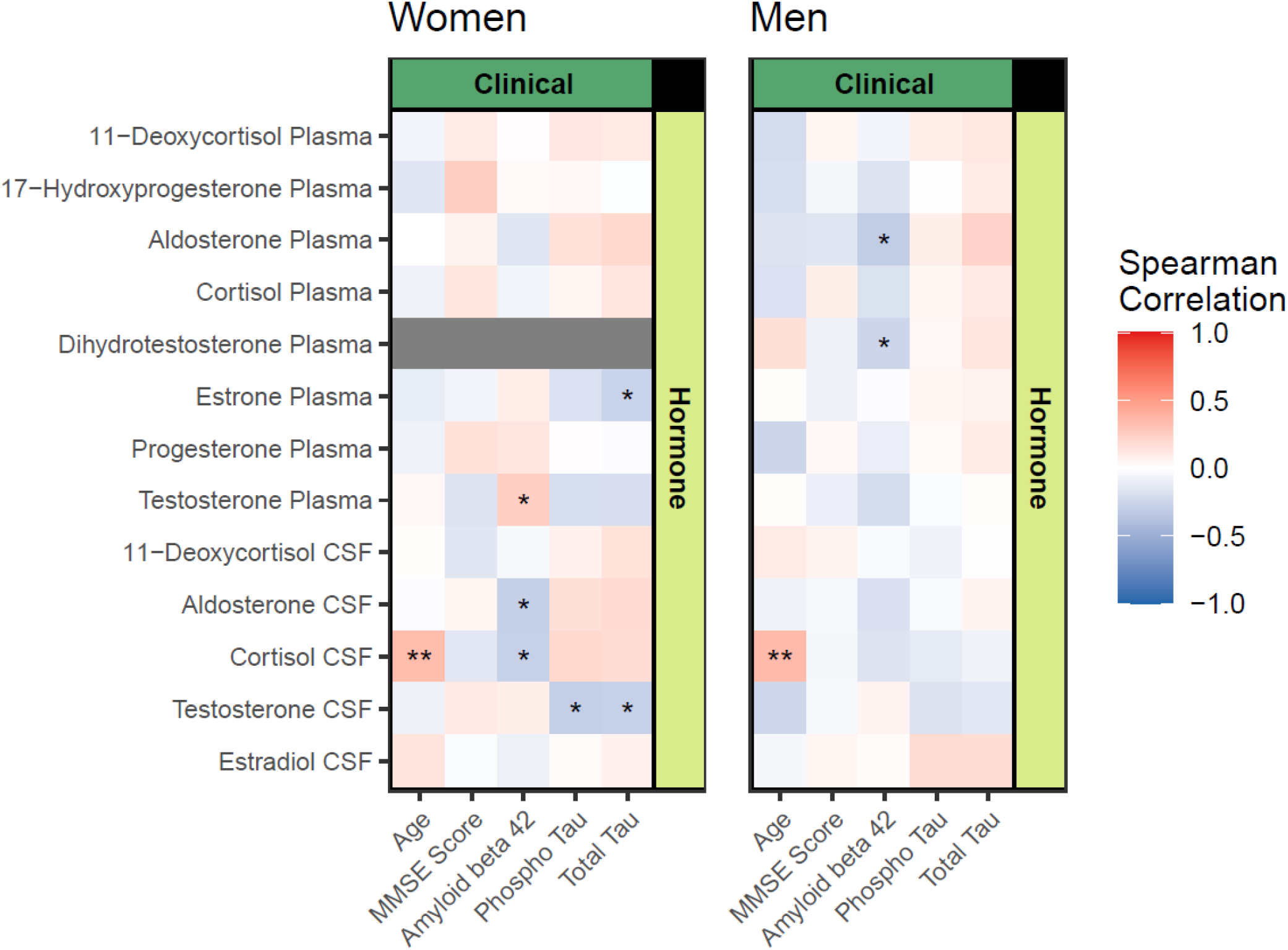
Correlation Matrix. Correlation Matrix: Heatmap of Spearman’s correlations between hormones and markers associated with AD. * indicate a q-value > 0.05, ** a q-value > 0.01 and *** a q-value > 0.001. q-values are obtained through multiple testing correction with FDR.

CSF cortisol increased with age in both sexes (women: r = 0.34, q = 0.01; men: r = 0.35, q = 1.9e-03; Supplementary table 2), whereas plasma cortisol showed no age association. In women, higher CSF cortisol was associated with lower Aβ-42 (r = −0.29, q = 0.01) and higher pTau (r = 0.20, p = 0.04), indicative of AD pathology. CSF cortisol in women also correlated positively with aldosterone in CSF and plasma, multiple plasma steroids (progesterone, testosterone, cortisol), and CSF 11-deoxycortisol. In men, CSF cortisol correlated with CSF and plasma aldosterone, plasma estrone, and CSF 11-deoxycortisol.

Aldosterone showed sex-specific associations with AD biomarkers: CSF aldosterone correlated negatively with Aβ-42 in women, whereas plasma aldosterone showed this association in men. Plasma aldosterone correlated positively with tTau in men but not women.

Testosterone displayed divergent sex-specific patterns. In women, CSF and plasma testosterone were negatively associated with pTau and tTau and positively associated with Aβ-42. In contrast, plasma testosterone in men correlated negatively with Aβ-42.

MMSE scores showed expected correlations with AD biomarkers, including positive associations with Aβ-42 and negative associations with tTau. However, significant correlations with hormones were limited, with only 17-hydroxyprogesterone showing a relationship with MMSE in women (r = 0.25, p-value = 0.01). Inter-hormone correlation patterns varied substantially by hormone class. Hormones detected and measured in both biofluids, including aldosterone, cortisol, 11-deoxycortisol, and testosterone, demonstrated strong correlations between their plasma and CSF concentrations. In contrast, estrogens showed minimal interconnectivity, plasma estrone and CSF estradiol correlated only with each other (negatively). At the opposite extreme, progesterone and 17-hydroxyprogesterone demonstrated the broadest correlation patterns, associating with all other plasma hormones except estrone.

Exploratory analyses using dimensionality reduction methods did not show any separation of the dementia subgroups in the unsupervised PCA (Supplementary Figure 5), nor for the supervised PLS-DA, yet the hormones with the most power to separate the dementia subgroups in CSF were cortisol, 11-deoxycortisol and aldosterone in both men and women subset. For plasma hormones the best separation power came from progesterone, 17-hydroxyprogesterone and testosterone for women and from 11-deoxycortisol, progesterone and 17-hydroxyprogesterone for men, showing some consistency across the sexes.

### Cortisol validation

Comparison of plasma cortisol measurements of 426 individuals from the ADNI cohort was carried out as an external validation. The cohort consisted of 153 cognitively healthy female participants, 133 cognitively healthy males, 56 female participants with AD and 84 male participants with AD. The mean normalized cortisol levels cognitively healthy female participants were 0.21, compared to 0.26 of the female participants with AD (log_2_ FC = 1.24, p-value = 1.79e-06, Figure 4, Supplementary table 3). The difference in men was less striking yet still significant, healthy male participants had a mean normalized cortisol level at 0.21 compared to male participants with AD 0.21 (log_2_ FC = 1.12, p-value = 0.04).

**Figure 4.**
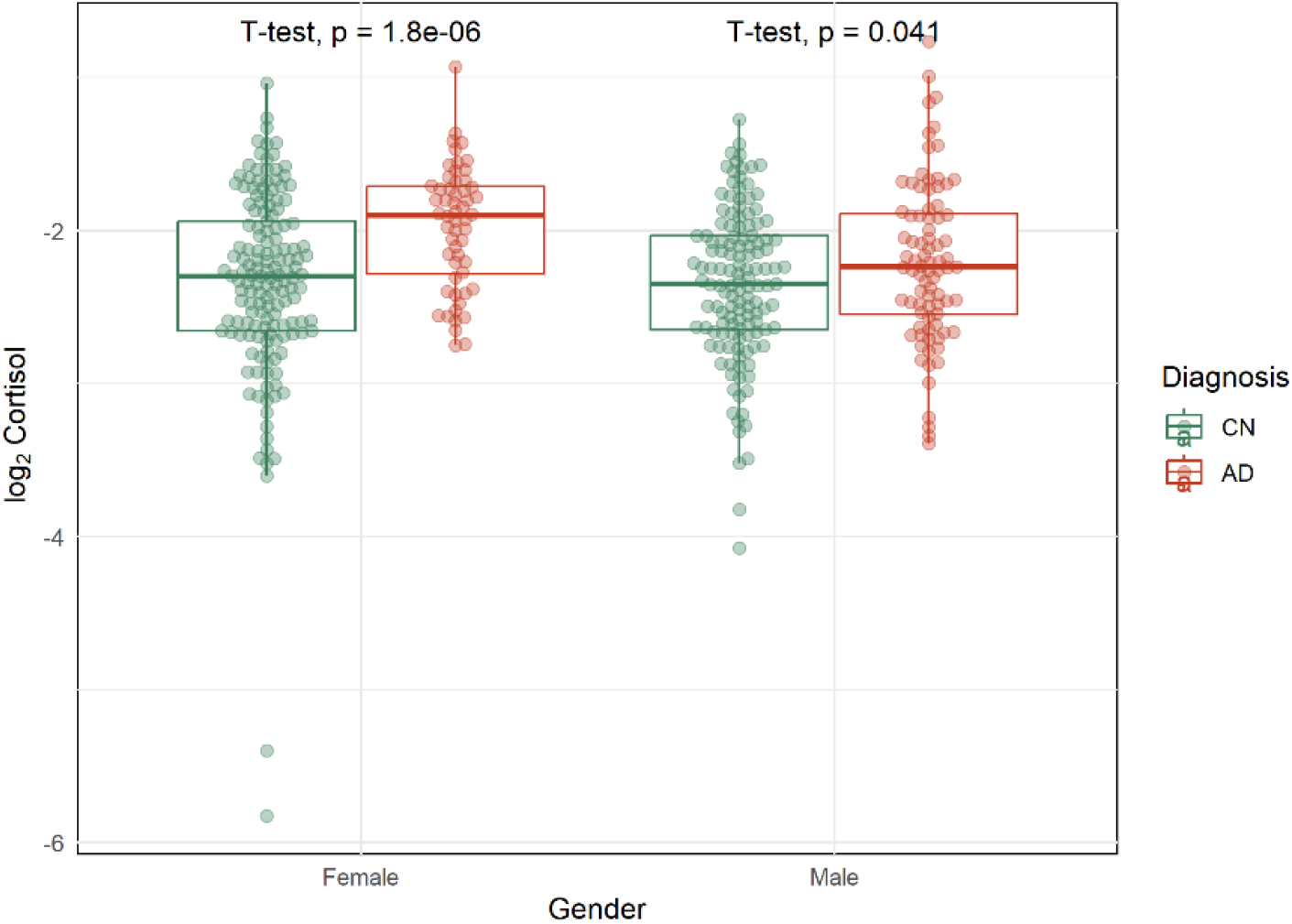
Differences between cortisol in ADNI. Boxplot with t-test comparison showing levels of cortisol in females and males with individuals thar are cognitively healthy (CN) or have Alzheimer’s disease (AD).

## Discussion

Here we have quantitatively measured hormone levels in both blood and CSF among older adults, addressing an important knowledge gap, particularly given the potential clinical implications for neurodegenerative diseases. For this we developed a method to quantify nine steroid hormones using mass spectrometry from CSF and plasma in a cohort of individuals in their 60s and 70s evaluated and diagnosed at the memory clinic at Rigshospitalet in Copenhagen. Our findings show significant sex specific associations between hormones and cognitive impairment specifically for cortisol, aldosterone and progesterone.

### CSF Cortisol and its precursor were higher in Women with AD

To our knowledge, this study is not only one of the few to measure cortisol in CSF but the only one to assess it both in CSF and plasma simultaneously. CSF cortisol and its precursor 11-deoxycortisol were significantly higher in women with AD (Figure 1), while no significant alterations were found in plasma or in men. A weaker trend toward higher CSF cortisol was observed in vascular dementia for both sexes. These results align with previous reports showing elevated plasma and CSF cortisol in AD and cognitive impairment ^16–20^. Additional testing of cortisol plasma from the ADNI cohort also showed increased levels of cortisol in people with AD compared to cognitively healthy individuals, this difference being larger in the female population (Figure 4). The reason we did not find any difference for plasma cortisol is likely due to the smaller sample size (n = 69 women vs. 203 women in ADNI, 44 men vs. 217 men in ADNI). Increased cortisol in AD has previously been shown in ADNI^25,26^, but here we highlight the difference between female and male participants.

While the lack of a plasma effect (FC (95% CI) = 1.01, (0.96-1.06), p-value = 0.7 in women) may reflect limited statistical power, it suggests that CSF cortisol (FC (95% CI) = 1.13, (1.01-1.27), p-value = 0.04) is a more sensitive marker of cognitive impairment than plasma cortisol. This is not surprising, as plasma cortisol is more readily influenced by circadian variation and external exposures, whereas CSF cortisol more directly reflects central hypothalamic– pituitary–adrenal (HPA) axis activity. Although CSF cortisol was highly correlated with age, our models were adjusted accordingly, suggesting that this association represents an independent effect.

Previous studies, such as the landmark study by Lupien *et al*., showed that elderly individuals with prolonged cortisol elevations over five years had reduced hippocampal volumes and memory deficits, with the degree of atrophy correlating strongly with cortisol levels ^27^. Elevated CSF cortisol was observed only in women, which may partly reflect limited statistical power (18 NCI and 51 women with AD vs. 14 NCI and 30 men with AD). However, there was little indication of a similar trend in men with AD, although a slight increase was observed in men with VaD. In women, CSF cortisol correlated negatively with Aβ-42 and positively with pTau, suggestive of AD pathology, correlations that were not seen in men (Figure 3). We have previously reported that AD presents distinct molecular manifestations in men and women ^28,29^. It has been suggested that estradiol decline after menopause exacerbates glucocorticoid sensitivity, leading to higher central cortisol exposure in women ^30^, however, we saw no correlation between estradiol and cortisol in this study.

The concurrent elevation of cortisol and its immediate precursor 11-deoxycortisol in CSF, in the absence of corresponding changes in plasma 17-hydroxyprogesterone and with decreased plasma progesterone, points to compartment-specific alterations in steroidogenesis within the brain/CSF. Since 11-deoxycortisol is converted to cortisol by 11β-hydroxylase, these findings suggest upstream local synthesis, upregulated enzymatic activity, or reduced clearance of cortisol precursors in the brain rather than systemic overproduction. Such central amplification could reflect neuroinflammatory modulation of steroidogenic enzymes or glial-driven neurosteroidogenesis, processes known to be altered in AD, particularly in women ^31^. Overall, our data highlight the potential of CSF cortisol and 11-deoxycortisol as a mechanism to study further in AD, especially among women.

### Aldosterone linked to brain pathology in men

We observed higher plasma aldosterone levels in men with AD, with a similar but non-significant trend in women (Figure 2), while CSF aldosterone showed no association. In men, plasma aldosterone correlated negatively with Aβ42 and positively with total tau, suggesting a possible link between peripheral mineralocorticoid activity and AD pathology (Figure 3). Previous studies have implicated dysregulation of the renin–angiotensin–aldosterone system (RAAS) in both AD and vascular dementia ^22,32^, and excessive aldosterone has been associated with cognitive impairment and reduced cerebral perfusion ^21,33^. Together, these findings suggest that elevated aldosterone may contribute to neurodegeneration indirectly through vascular or inflammatory pathways rather than direct central accumulation, consistent with the lack of association in CSF. The stronger effect in men may reflect sex differences in RAAS regulation or vascular susceptibility.

### Progesterone, Estrogens and Testosterone

Plasma progesterone concentrations were significantly reduced in women with AD (Figure 2), as well as across all disease categories (MCI-AD, AD, and VaD) in our combined models (Supplementary figure 2). Hydroxy-progesterone showed a similar but less pronounced decrease, it was reduced significantly in vascular dementia suggesting a shift towards mineralocorticoid synthesis.

Progesterone receptors are broadly expressed throughout the brain, in key cognitive regions including the hippocampus and frontal cortex, with no apparent restriction to specific cell types or hormonal status ^34^. Progesterone also exerts important protective effects on the cardiovascular system that could be relevant to vascular health. The natural hormone acts as a vasodilator through modulation of calcium channels and enhancement of nitric oxide bioavailability, improving cerebral blood flow ^35,36^.

While progesterone supplementation could represent a potential therapeutic strategy, implementing such treatment is far from straightforward. Progesterone can be found as different formulations, as an example, progestins, a range of progesterone derivatives commonly used for birth control and in hormone replacement therapy (HRT), may act differently than endogenous progesterone ^37,38^. Some studies have also reported higher dementia risk associated with HRT use, highlighting that we need more studies ^39,40^. The specific molecular composition of HRT formulations would be a critical factor in any prevention trials together with choosing the appropriate therapeutic window and dose.

A meta-analysis by Rocca *et al*. demonstrated that HRT may be detrimental for women in late post-menopause, specifically the age of the participant in our study (65-79), while conferring neuroprotective effects in early post-menopause (ages 50-60) ^41^. This time-dependent effect has been confirmed in subsequent studies ^7,8,42^. One study found HRT improved memory delay and associated with larger brain volumes but only in women carrying the APOE4 genotype ^43^.

We observed decreased CSF estradiol in women with VaD (FC (95% CI) = 0.86, (0.74-0.98), p-value = 0.03), which interestingly was not seen in the AD group (FC (95% CI) = 1.03, (0.93-1.14), p-value = 0.6). In plasma, higher estrone was associated with VaD in men and showed a similar trend in MCI-AD and AD. These findings support sex-specific alterations in estrogenic pathways across dementias. Reduced estradiol in women could contribute to vascular vulnerability, consistent with epidemiological studies linking shorter lifetime estrogen exposure or early menopause to greater dementia risk ^5,6^. One study of the UK biobank found no causal link between estradiol exposure and brain health, suggesting that associations between estradiol and brain health is governed by hormone fluctuations ^44^. Estrogens exert neuroprotective effects through modulation of synaptic plasticity, cerebral blood flow, and amyloid metabolism ^45^. In men, however, elevated estrone may reflect compensatory aromatization of testosterone under metabolic or vascular stress, possibly signalling altered peripheral-to-central steroid dynamics rather than direct neuroprotection.

No overall association was found between testosterone and cognitive decline, but sex-specific patterns emerged. In women, CSF and plasma testosterone correlated negatively with tau markers and positively with Aβ42, suggesting a protective role. In men, plasma testosterone correlated negatively with Aβ42, consistent with prior studies linking low testosterone to higher dementia risk ^14,15,46^. These divergent associations likely reflect sex differences in androgen metabolism and receptor sensitivity, emphasizing that hormonal ratio balance, rather than absolute levels, may be key to maintaining cognitive health.

### Strengths and limitations

To our best knowledge this is the first comprehensive study to fully quantify aldosterone, 11-deoxycortisol, cortisol, and testosterone in both plasma and CSF, with a total of nine unique hormones across the two biofluids, from the same elderly individuals across diagnosis. Five distinct dementia subgroups were defined, based on a thorough clinical evaluation in the memory clinic. Some limitations are the cross-sectional design of this study, hormone measurements include a single time-point and therefore they cannot capture hormone dynamics that will be important for AD risk. As the persons in this study are considered vulnerable, samples collection did not follow a fasting period. Additionally, the stratification of participants by both dementia subtype and sex resulted in relatively small group sizes, which may have limited our statistical power.

### Conclusion

In summary, this study demonstrates distinct, sex-specific associations between steroid hormones and cognitive impairment. Women exhibited elevated central cortisol and reduced peripheral progesterone, whereas men showed higher peripheral aldosterone. These findings suggest that dysregulation of glucocorticoid, mineralocorticoid, and sex-steroid pathways could contribute to neurodegeneration through both central and vascular mechanisms. The divergent hormone patterns underscore the need for sex-specific considerations in AD and related dementias research.

## Data Availability

All data produced in the present study are available upon reasonable request to the authors

## List of abbreviations

Abbreviation Full term

AD: Alzheimer’s disease
ADNI: Alzheimer’s Disease Neuroimaging Initiative
APOEε4: Apolipoprotein E ε4
Aβ-42: Amyloid-β 42
CI: Confidence interval
NC: Cognitively healthy control
CSF: Cerebrospinal fluid
dMRM: Dynamic multiple reaction monitoring
FC: Fold change
GLM: Generalized linear model
LC-MS: Liquid chromatography–tandem mass spectrometry
MCI: Mild cognitive impairment
NCI: No cognitive impairment
PCA: Principal component analysis
LPS-DA: Partial least squares–discriminant analysis
pTau: Phosphorylated tau
SEM: Standard error of the mean
tTau: Total tau
VIP: Variable importance in projection

## Statements

## Acknowledgement

We would like to acknowledge the participants from the Danish Dementia Center at Copenhagen University Hospital, Rigshospitalet for making this study possible.

## Author Contribution

Contributions to this article follows the Contributor Role Taxonomy (CRediT). **T.M**.: Conceptualization, Formal analysis, Methodology, Project administration, Software, Visualization, Writing - original draft, Writing - review & editing. **A.W**.: Conceptualization, Formal analysis, Methodology, Project administration, Software, Visualization, Writing - original draft, Writing - review & editing. **K.H**.: Formal analysis, Methodology, Project administration, Writing - review & editing. **M.C.-G**.: Formal analysis, Methodology, Software, Visualization, Writing - review & editing. **L.Y**.: Formal analysis, Methodology, Software, Visualization, Writing - review & editing. **A.H.S**.: Data curation, Resources, Writing - review & editing. **L.W**.: Writing - review & editing. **T.S.A:** Writing - review & editing. **P.P**.: Writing - review & editing. **R.E.M**.: Funding acquisition, Writing - review & editing. **T.K**.: Writing - review & editing. **S.G.H**.: Funding acquisition, Project administration, Data curation, Resources, Writing - review & editing. **C.L.-Q**.: Conceptualization, Funding acquisition, Project administration, Supervision, Writing – original draft, review & editing.

## Availability of data and materials

The dataset analyzed here is not directly publicly available, for the privacy of the participants, in compliance with EU and Danish data protection law. The data can be accessed upon reasonable request and relevant legal permission from the danish data protection agency. Data access requests should be directed to S.G.H..

## Declaration of interest

T.M., A.W., K.H., M.C.-G., L.Y., L.W., P.P., R.E.M., T.K, S.G.H., C.L.-Q. declare no conflicts of interest.

A.H.S. has received a one-time consulting fee from Eisai / BioArctic, paid to the institution (2025). T.S.A. owns stocks in NovoNordisk and Zealand Pharma.

## Ethical approval

Protocols and procedures were approved by the relevant local ethical committees.

## Source of funding

Funding for this work was provided by Lundbeck Fonden by grant R344-2020-989.

## Notes

### Author Declarations

Approved by the Danish Health Research Ethics Committee B for the capital Region - H-21051757

## References

1. Rajan KB, Weuve J, Barnes LL, et al. Population estimate of people with clinical Alzheimer’s disease and mild cognitive impairment in the United States (2020–2060). Alzheimer’s Dement. 2021;17(12):1966–1975. doi:10.1002/alz.12362

2. Matthews FE, Stephan BCM, Robinson L, et al. A two decade dementia incidence comparison from the Cognitive Function and Ageing Studies i and II. Nat Commun. 2016;7. doi:10.1038/ncomms11398

3. Beam CR, Kaneshiro C, Jang JY, Reynolds CA, Pedersen NL, Gatz M. Differences between Women and Men in Incidence Rates of Dementia and Alzheimer’s Disease. J Alzheimer’s Dis. 2018;64(4):1077–1083. doi:10.3233/JAD-180141

4. Zhao Y, Wang Q, Fu C, et al. Sex Hormones and Risk of Incident Dementia in Men and Postmenopausal Women. Clin Endocrinol (Oxf). 2025;103:366–375. doi:10.1111/cen.15271

5. Id JG, Harris K, Peters SAE, et al. Reproductive factors and the risk of incident dementia: A cohort study of UK Biobank participants. PLoS Med. 2022;18:1–23. doi:10.1371/journal.pmed.1003955

6. Liao H, Cheng J, Pan D, et al. Articles Association of earlier age at menopause with risk of incident dementia, brain structural indices and the potential mediators: a prospective community-based cohort study. eClinicalMedicine. 2023;60:1–14. doi:10.1016/j.eclinm.2023.102033

7. Matyi JM, Rattinger GB, Schwartz S, Buhusi M, Tschanz JT. Lifetime estrogen exposure and cognition in late life: the Cache County Study. Menopause J ofThe North Am Menopause Soc. 2019;26(12):1366–1374. doi:10.1097/GME.0000000000001405

8. Coughlan GT, Betthauser TJ, Boyle R, et al. Association of Age at Menopause and Hormone Therapy Use With Tau and β -Amyloid Positron Emission Tomography. JAMA Neurol. 2023;80(5):462–473. doi:10.1001/jamaneurol.2023.0455

9. Park HK, Sc M, Ph D, et al. The Effects of Estrogen on the Risk of Developing Dementia: A Cohort Study Using the UK Biobank Data. Am J Geriatr Psychiatry. Published online 2024:1–14. doi:10.1016/j.jagp.2024.01.025

10. Hao W, Fu C, Dong C, et al. Age at menopause and all-cause and cause-specific dementia: a prospective analysis of the UK Biobank cohort. Hum Reprod. 2023;38(9):1746–1754.

11. Lange A marie G De, Barth C, Maximov II, et al. Women’s brain aging: Effects of sex-hormone exposure, pregnancies, and genetic risk for Alzheimer’s disease. Hum Brain Mapp. 2020;41(March):5141–5150. doi:10.1002/hbm.25180

12. Najar J, Hallstrom T, Zettergren A, et al. Reproductive period and preclinical cerebrospinal fluid markers for Alzheimer disease: a 25-year study. Menopause J ofThe North Am Menopause Soc. 2021;28(10):1099–1107. doi:10.1097/GME.0000000000001816

13. Zhang Z, Kang D, Li H. Testosterone and Cognitive Impairment or Dementia in Middle-Aged or Aging Males : Causation and Intervention, a Systematic Review and Meta-Analysis. J Geriatr Psychiatry Neurol. 2021;34(1):405–417. doi:10.1177/0891988720933351

14. Marriott RJ, Murray K, Flicker L, et al. Lower serum testosterone concentrations are associated with a higher incidence of dementia in men: The UK Biobank prospective cohort study. Alzheimer’s Dement. 2022;18(August 2021):1907–1918. doi:10.1002/alz.12529

15. Yeung CHC, Yeung SLA, Kwok MK, Zhao J V., Schooling CM. The influence of growth and sex hormones on risk of alzheimer ‘ s disease : a mendelian randomization study. Neuro-Epidemiology. 2023;38:745–755.

16. Peña-bautista C, Baquero M, Ferrer I, Hervás D, Vento M, García-blanco A. Neuropsychological assessment and cortisol levels in biofluids from early Alzheimer’s disease patients. Exp Gerontol. 2019;123(May):10–16. doi:10.1016/j.exger.2019.05.007

17. Zheng B, Tal R, Yang Z, Middleton L, Udeh-momoh C. Cortisol hypersecretion and the risk of Alzheimer’s disease: A systematic review and meta-analysis. Ageing Res Rev J. 2020;64(March).

18. Ennis GE, Resnick SM, Ferrucci L, Brien RJO. Long-term cortisol measures predict Alzheimer disease risk. Neurology. 2016;88:371–378. doi:10.1212/WNL.0000000000003537

19. Pietrzak RH, Laws SM, Lim YY, et al. Archival Report Plasma Cortisol, Brain Amyloid-β, and Cognitive Decline in Preclinical Alzheimer’s Disease: A 6-Year Prospective Cohort Study. Biol Psychiatry Cogn Neurosci Neuroimaging. 2017;2(January):45–52. doi:10.1016/j.bpsc.2016.08.006

20. Udeh-momoh CT, Su B, Evans S, Zheng B, Sindi S, Sciences H. Cortisol, Amyloid-β, and Reserve Predicts Alzheimer’s Disease Progression for Cognitively Normal Older Adults. J Alzheimer’s Dis. 2019;70:553–562. doi:10.3233/JAD-181030

21. Yagi S, Akaike M, Aihara K ichi, et al. High plasma aldosterone concentration is a novel risk factor of cognitive impairment in patients with hypertension. Hypertens Res. 2011;(June 2010):74–78. doi:10.1038/hr.2010.179

22. Hong N, Kim KJ, Yu MH, Jeong SH, Lee S, Lim JS. Risk of dementia in primary aldosteronism compared with essential hypertension : a nationwide cohort study. Alzheimers Res Ther. Published online 2023:1–12. doi:10.1186/s13195-023-01274-x

23. Hasselbalch SG, Andersen BB, Ejlerskov P, et al. The Danish Dementia Research Centre: Integrating patient care, clinical research, and national educational services. J Alzheimer’s Dis. 2025;107(3):883–898. doi:10.1177/13872877251365195

24. Mueller SG, Weiner MW, Thal LJ, et al. Ways toward an early diagnosis in Alzheimer’s disease: The Alzheimer’s Disease Neuroimaging Initiative (ADNI). Alzheimer’s Dement. 2005;1:55–66. doi:10.1016/j.jalz.2005.06.003

25. Toledo JB, Weiner MW, Jack CR. Cardiovascular risk factors, cortisol, and amyloid-b deposition in Alzheimer’s Disease Neuroimaging Initiative. Alzheimer’s Dement. 2012;8:483–489. doi:10.1016/j.jalz.2011.08.008

26. Borkowski K, Yin C, Kindt A, et al. Metabolic Alteration in Oxylipins and Endocannabinoids Point to an Important Role for Soluble Epoxide Hydrolase and Inflammation in Alzheimer’s Disease - Finding from Alzheimer’s Disease Neuroimaging. BioRxiv. Published online 2025:1–28.

27. Lupien SJ, Leon M De, Santi S De, et al. Cortisol levels during human aging predict hippocampal atrophy and memory deficits. Nat Neurosci. 1998;1(1):69–73.

28. Xu J, Bankov G, Kim M, et al. Integrated lipidomics and proteomics network analysis highlights lipid and immunity pathways associated with Alzheimer’s disease. Transl Neurodegener. 2020;9(1):1–15.

29. Wretlind A, Xu J, Chen W, et al. Lipid profiling reveals unsaturated lipid reduction in women with Alzheimer’s disease. Alzheimer’s Dement. 2025;21(8):1–13. doi:10.1002/alz.70512

30. Cohn AY, Grant LK, Nathan MD, et al. Effects of Sleep Fragmentation and Estradiol Decline on Cortisol in a Human Experimental Model of Menopause. J Clin Endocrinol Metab. 2023;108(11):1347–1357. doi:10.1210/clinem/dgad285

31. Klyubin I, Ondrejcak T, Hu N wei, Rowan MJ. ScienceDirect Glucocorticoids, synaptic plasticity and Alzheimer ‘ s disease. Curr Opin Endocr Metab Res. 2022;25:100365. doi:10.1016/j.coemr.2022.100365

32. Tayler HM, Kehoe PG, Miners JS. Dysregulation of the renin-angiotensin system in vascular dementia. Brain Pathol. 2024;34(October 2023):1–12. doi:10.1111/bpa.13251

33. Hajjar I, Hart M, Mack W, Lipsitz LA. Aldosterone, Cognitive Function, and Cerebral Hemodynamics in Hypertension and Antihypertensive Therapy. Am J Hypertens. 2015;28(March):319–325. doi:10.1093/ajh/hpu161

34. Brinton RD, Thompson RF, Foy MR, et al. Progesterone receptors : Form and function in brain. Front Neuroendocrinol. 2008;29:313–339. doi:10.1016/j.yfrne.2008.02.001

35. Oelkers WKH. Effects of estrogens and progestogens on the renin-aldosterone system and blood pressure. Steroids. 1996;(96):166–171.

36. Hermenegildo C, Oviedo PJ, Garcia-Martinez MC, Garcia-Perez MA, Tarin JJ, Cano A. Progestogens stimulate prostacyclin production by human endothelial cells. Hum Reprod. 2005;20(6):1554–1561. doi:10.1093/humrep/deh803

37. Cordina-duverger E, Truong T, Anger A, et al. Risk of Breast Cancer by Type of Menopausal Hormone Therapy: a Case-Control Study among Post-Menopausal Women in France. PLoS One. 2013;8(11):1–9. doi:10.1371/journal.pone.0078016

38. Asi N, Mohammed K, Haydour Q, et al. Progesterone vs. synthetic progestins and the risk of breast cancer: a systematic review and meta-analysis. Syst Rev. 2016;5(121):1–8. doi:10.1186/s13643-016-0294-5

39. Savolainen-peltonen H, Rahkola-soisalo P, Hoti F, et al. Use of postmenopausal hormone therapy and risk of Alzheimer’s disease in Finland: nationwide case-control study. BMJ. 2019;364:1–8. doi:10.1136/bmj.l665

40. Pourhadi N, Mørch LS, Holm EA, Torp-pedersen C, Meaidi A. Menopausal hormone therapy and dementia: nationwide, nested case-control study. BMJ. Published online 2023:1–7. doi:10.1136/bmj-2022-072770

41. Rocca WA, Grossardt BR, Shuster LT. Molecular and Cellular Endocrinology Oophorectomy, estrogen, and dementia: A 2014 update. Mol Cell Endocrinol. 2014;389:7–12. doi:10.1016/j.mce.2014.01.020

42. Song Y jia, Li S ran, Li X wan, et al. The Effect of Estrogen Replacement Therapy on Alzheimer’s Disease and Parkinson’s Disease in Postmenopausal Women: A Meta-Analysis. Front Neurosci. 2020;14(March):1–13. doi:10.3389/fnins.2020.00157

43. Saleh RNM, Hornberger M, Ritchie CW, Minihane AM. Hormone replacement therapy is associated with improved cognition and larger brain volumes in at-risk APOE4 women: results from the European Prevention of Alzheimer’s Disease (EPAD) cohort. Alzheimer’s Res Ther. 2023;15(1):1–13. doi:10.1186/s13195-022-01121-5

44. Oppenheimer H, Meer D van der, Schindler LS, et al. No causal links between estradiol and female’s brain and mental health using Mendelian randomization. Nat Commun. 2025;16(10915):1–13.

45. Kim G won, Park K, Jeong G woo. Effects of Sex Hormones and Age on Brain Volume in Post-Menopausal Women. J Sex Med. 2018;15(5):662–670. doi:10.1016/j.jsxm.2018.03.006

46. Liu L, Zhang C, Lv X, et al. Sex-specific associations between lipids and cognitive decline in the middle-aged and elderly: a cohort study of Chinese adults. Alzheimer’s Res Ther. 2020;12(1):1–13. doi:10.1186/s13195-020-00731-1

