## Supplementary material and figures for "Steroid Hormones in Dementia: A Cross-Diagnostic Molecular Analysis of Blood and Cerebrospinal Fluid"

### Index

**S1. – Hormone quantification method**

Sample Preparation and Extraction

Steroid hormone analysis was performed using identical extraction procedures for both plasma and cerebrospinal fluid (CSF) samples. Briefly, 50 µL of plasma or CSF was added to 1.5-2 mL Eppendorf tubes, followed by the addition of 440 µL acetone:acetonitrile (1:1 v/v) for protein precipitation and analyte extraction. The mixture was vortexed for 10 seconds and centrifuged at 10,000 × g for 5 minutes at 4°C. Subsequently, 450 µL of supernatant was transferred to clean tubes and dried using a speed vacuum system. The dried residue was reconstituted in 50 µL methanol:water (1:1 v/v) and transferred to amber glass LC vials with inserts. Samples were stored at -80°C until analysis.

Liquid Chromatography-Mass Spectrometry Analysis

The analytical system comprised an Agilent 1290 Infinity UHPLC system coupled with an Agilent 6460 triple quadrupole mass spectrometer (Agilent Technologies Inc., Santa Clara, CA, USA). Chromatographic separation was achieved using a Waters HSS T3 column (1.8 µm, 2.1 × 100 mm) protected by a C18 HSS T3 VanGuard pre-column (100Å, 1.8 µm, 2.1 mm × 5 mm). The mobile phase consisted of eluent A (water with 2 mM ammonium fluoride) and eluent B (methanol without modifier) at a flow rate of 0.4 mL/min. The gradient program was as follows: 0-1 min, 20% B; 1-2 min, linear gradient to 80% B; 2-8 min, linear gradient to 100% B; 8-9 min, 100% B; 9-9.2 min, return to 20% B; 9.2-12 min, 20% B for column re-equilibration.

Mass Spectrometry Detection

Mass spectrometric detection was performed using electrospray ionization (ESI) in positive mode with dynamic multiple reaction monitoring (dMRM). Instrument-dependent parameters were set as follows: nitrogen drying gas flow rate, 12 L/min at 325°C; capillary voltage, 3500 V; nebulizer pressure, 45 psi; nitrogen sheath gas flow rate, 11.0 L/min at 325°C; cell accelerator voltage, 4 V. Specific MRM transitions, retention times, fragmentor voltages, and collision energies for each hormone are provided in Supplementary Table 1.

Quality Control and Quantification

Each hormone was quantified using external calibration curves with specific standards. Two sets of 21-point calibration curves were prepared, ranging from 0.00075 to 1000 ppb (0.00075, 0.0015, 0.003, 0.006, 0.0121, 0.0243, 0.048, 0.0975, 0.195, 0.39, 0.78, 1.56, 3.125, 6.25, 12.5, 25, 50, 100, 200, 400, 1000 ppb), with one set analyzed at the beginning and one at the end of each analytical run. Quality control included two blank samples and one reference pooled sample analyzed for every 10 study samples. Data processing and quantification were performed using Agilent MassHunter Quantitative Analysis Software (version 10.2).

**S1. Table – Ion fragmentation conditions**

| **Hormone** | **Precursor ion** | **Product ion** | **Retention time** | **Fragmentor** | **Collision Energy** |
|---|---|---|---|---|---|
| 11-Deoxycortison | 347.2 | 109 | 3.3 | 150 | 29 |
| 17-Hydroxyprogesterone | 331.2 | 109 | 3.66 | 128 | 33 |
| Aldosterone | 361.2 | 342.2 | 3 | 150 | 13 |
| Cortisol | 363.3 | 121 | 3.15 | 128 | 25 |
| Dihydrotestosterone | 291.2 | 255 | 3.95 | 150 | 13 |
| Estrone | 271.1 | 253.1 | 3.5 | 150 | 13 |
| Progesterone | 315.2 | 109 | 4.16 | 144 | 25 |
| Testosterone | 289.2 | 109 | 3.6 | 128 | 25 |
| Estradiol | 271.1 | 183 | 3.4 | 128 | 49 |

**Supplementary Figure 1 – CSF GLM (all)**

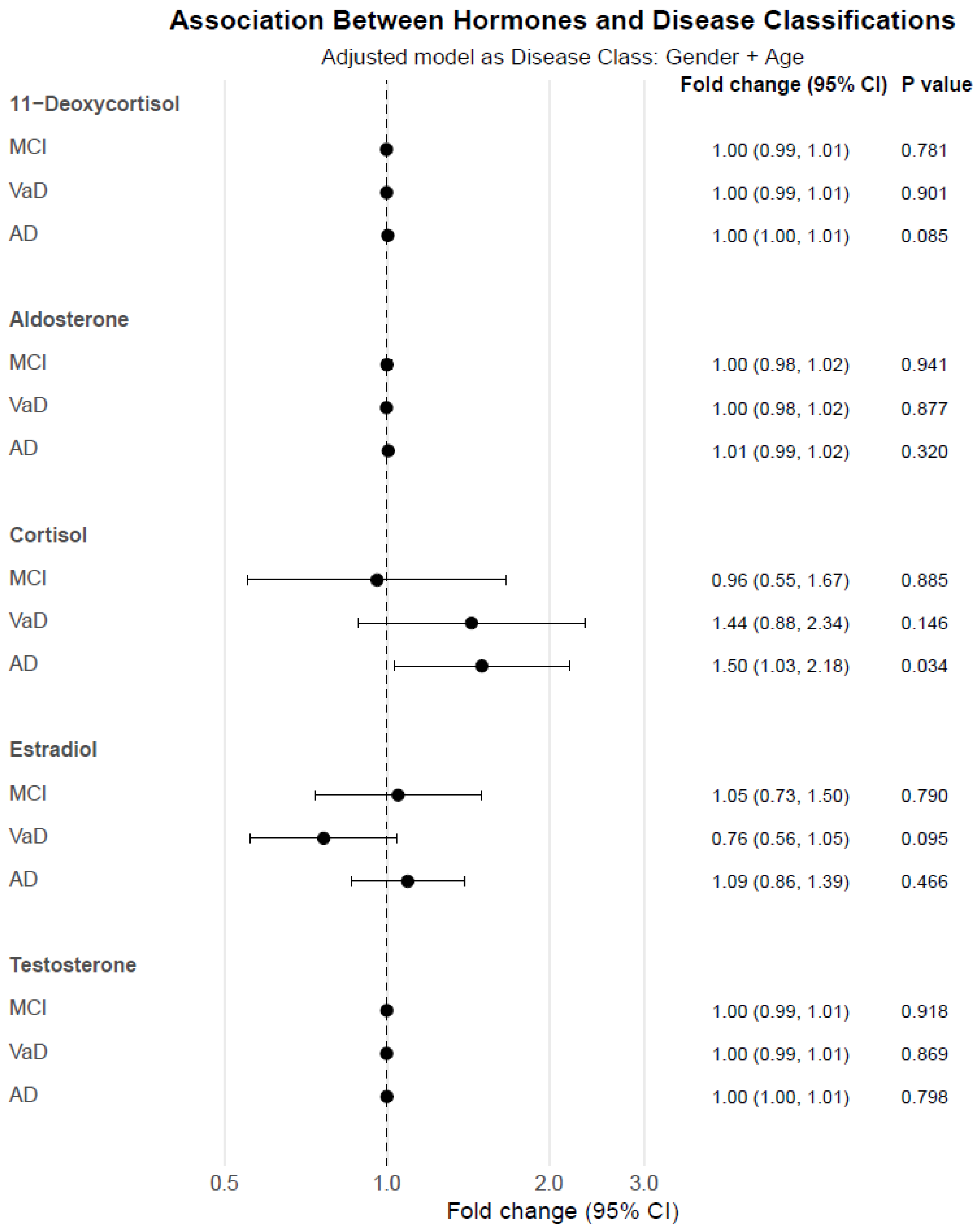

Supplementary Figure 1: Association between CSF hormones and disease classification using GLM. Fold change with 95% confidence interval against NCI and p-values are shown on the right.

**Supplementary Figure 2 – Plasma GLM (all)**

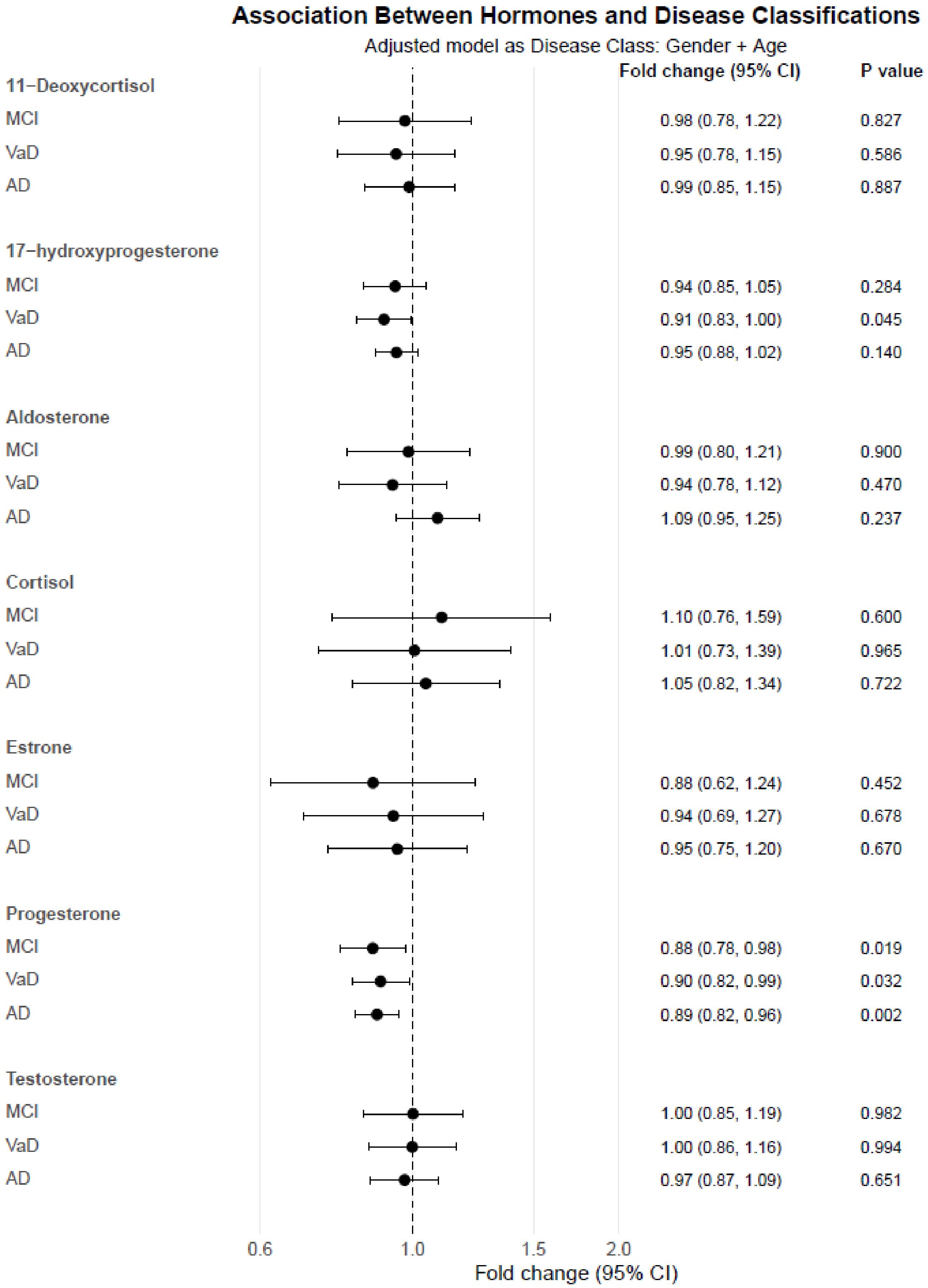

Supplementary Figure 2: Association between plasma hormones and disease classification using GLM. Fold change with 95% confidence interval against NCI and p-values are shown on the right.

**Supplementary Figure 3 – Correlations Full (sex stratified)**

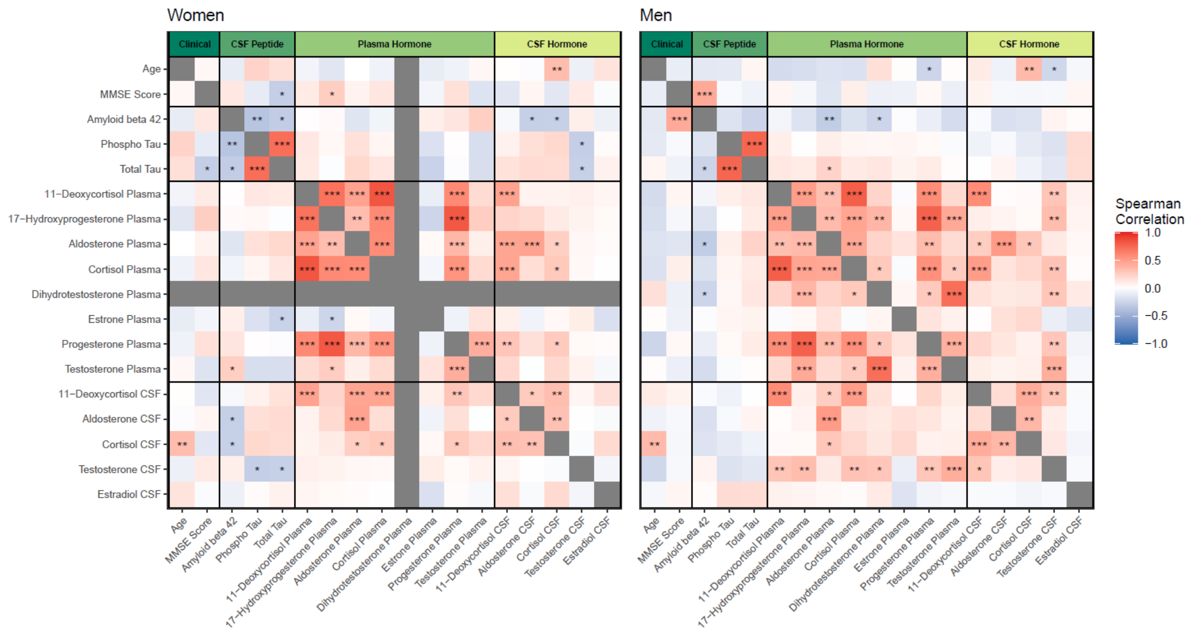

Supplementary Figure 3: Correlation matrix: Heatmap of Spearman's correlations between hormones and markers associated with AD. Data is separated by sex with women on the left and men on the right. * indicate a q-value > 0.05, ** a q-value > 0.01 and *** a q-value > 0.001. q-values are obtained through multiple testing correction with FDR.

**Supplementary Figure 4 – Correlations Full (all)**

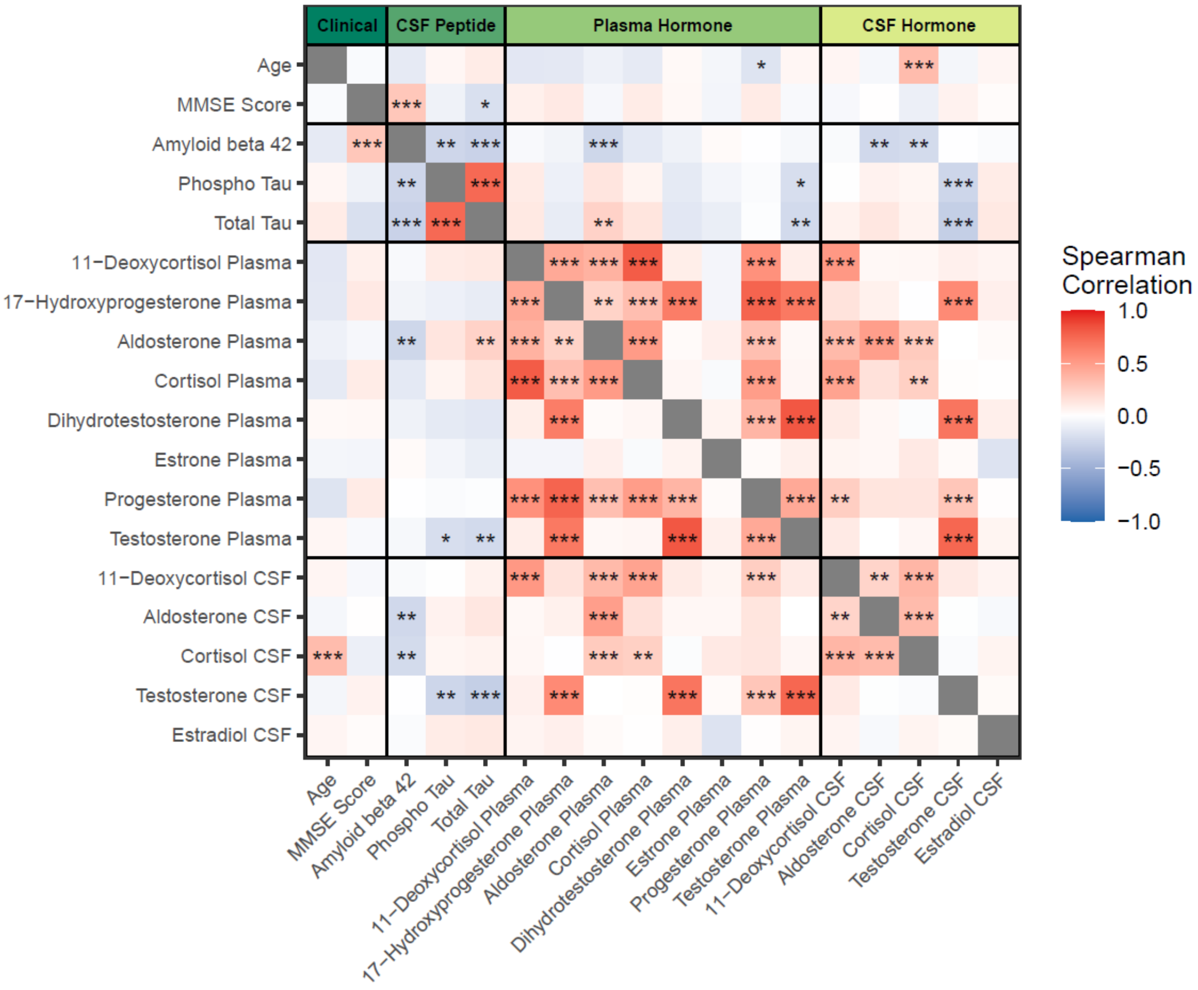

Supplementary Figure 4: Correlation matrix: Heatmap of Spearman's correlations between hormones and markers associated with AD. * indicate a q-value > 0.05, ** a q-value > 0.01 and *** a q-value > 0.001. q-values are obtained through multiple testing correction with FDR.

### Supplementary Figure 5 – PCA and PLS-DA

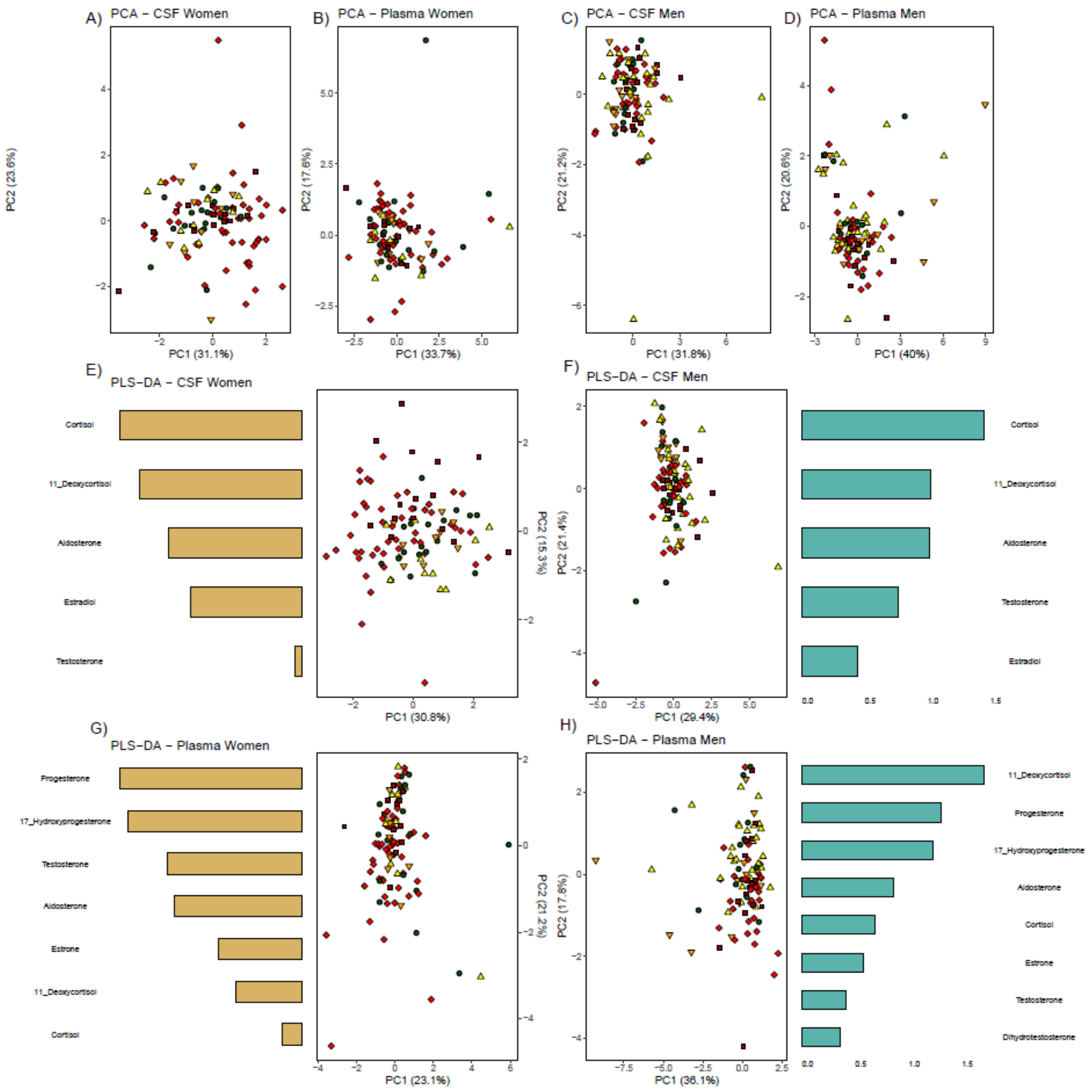

Supplementary Figure 5: PCA and PLS-DA: PCA plots of hormone data in different subsets are visualized at the top, CSF in women, plasma in women, CSF men, and plasma men shown A-D respectively. ● = No cognitive impairment, ▲ = MCI_Non-AD, ▼ = MCI, ■ = VaD, ♦ = AD. E-F show PLS-DA plots of hormone data in the same subsets, women on the left and men on the right. Disease class is used as separator, next to each PLS-DA plot is the five hormones with the highest VIP score in the separation.
